# A protocol for the TRACS-Liverpool study, tracking transmission of extended-spectrum beta-lactamase producing Enterobacterales across health and social care settings in the United Kingdom

**DOI:** 10.64898/2026.05.13.26352872

**Authors:** Sarah Gallichan, Joseph M Lewis, Sally Forrest, Maria Moore, Esther Picton-Barlow, Claudia McKeown, Christopher P. Jewell, Stacy Todd, Fabrice E Graf, Nicholas A Feasey

## Abstract

**Background:** Antimicrobial resistance (AMR) is a global public health problem. Infections caused by extended-spectrum beta-lactamase (ESBL) and carbapenemase (CP) - producing Enterobacterales (E) threaten individuals and healthcare systems worldwide. Symptomatic infection caused by Enterobacterales is typically preceded by asymptomatic colonisation and often occurs in the most vulnerable individuals, thus interrupting asymptomatic transmission is desirable. The dominant transmission routes across the healthcare continuum including hospitals, intermediate care, and long-term care facilities are not well understood.

**Methods:** Here we present a protocol describing a genomic surveillance framework developed for the *Tracking Antimicrobial Resistance Across Care Settings (TRACS) Liverpool* programme, which aims to identify critical ESBL-E transmission points in hospitals and care homes in Liverpool, UK. Our study integrates individual participant and healthcare facility data, validated standard operating procedures for taking and culturing stool, rectal, environmental, and staff samples, and genomic sequencing of ESBL-E, and statistical modelling approaches into a research framework for ESBL-E genomic surveillance.

**Discussion:** There is a need for improved epidemiological and laboratory approaches to studying bacterial transmission. Drug-resistant enteric bacteria are a highly tractable marker of the movement of all enteric bacteria, and interventions designed to interrupt transmission of drug-resistant bacteria are expected to have a broader healthcare impact. This protocol provides a standardised, reproducible approach for identifying ESBL-E, tracking acquisition events, and linking clinical and environmental isolates through whole-genome sequencing.

## Background

Antimicrobial resistance (AMR) remains a critical global health challenge with profound implications for morbidity, mortality, and healthcare system sustainability^1^. Extended-spectrum beta-lactamase (ESBL) and carbapenemase (CP) -producing Enterobacterales (E) are particularly concerning due to their ability to colonise hosts asymptomatically^2^, persist in built environments^3^, and evade multiple classes of antibiotics. Accordingly, ESBL-E and CPE are of critical priority on WHO’s (World Health Organisation) bacterial priority pathogens list^4^.

ESBL-E and CPE organisms thrive in complex care systems characterised by vulnerable populations, frequent antibiotic exposure, and interconnected patient pathways. Elderly people moving between care facilities and healthcare environments are well known to be at considerable risk of ESBL-E and CPE infections in high-income countries^1,5^. In the UK, residential and nursing care is largely private sector led, especially in Liverpool, where the local authority operates no care homes. Average weekly fees in the UK for residential care are £1298 and £1535 for nursing care^6^, however, facilities and standards vary widely. Funding options also vary, while some residents are eligible for funding from the local authority or, in some cases, the National Health Service (NHS), most residents are self-funded.

Both care homes and hospitals are inspected by the Care Quality Commission, the independent regulator for health and social care. Inspection criteria include infection prevention and control (IPC) measures; however, these are resource-intensive to routinely assess and the extent to which IPC programmes are robustly assessed as part of this process is not clear. Asymptomatic colonisation by enteric bacteria typically precedes symptomatic infection and whilst frailer individuals are typically at greater risk of colonisation progressing to infection, it is not easy to predict who will or will not develop a mild, moderate or severe, life-threatening infection. We therefore hypothesise, that health systems should aim to interrupt asymptomatic transmission and thus prevent infection.

Multiple studies have documented the role of environmental reservoirs such as toilets, sinks, U-bends, drains, and high-touch surfaces in hospitals in propagating drug-resistant organisms^7–9^. However, the relative contribution of environmental contamination, patient-to-patient transmission, and staff-mediated spread in long-term healthcare facilities is poorly resolved. The lack of standardised sampling methodologies, inconsistent inclusion of environmental sampling, and lack of integrated genomic analysis aimed at distinguishing between different bacterial populations of the same bacterial species within complex samples have limited our ability to draw definitive conclusions. Further, it is quite possible that the majority of Enterobacterales implicated in healthcare associated infection are acquired in the community^10^. The COVID-19 pandemic illustrated the potential for respiratory transmission of pathogens in long-term care facilities^11^, but the potential for transmission of enteric bacteria implicated in the “silent pandemic” of AMR in long-term care facilities is under explored.

It is important to understand and thus propose measures to prevent the spread of ESBL-E and CPE throughout the care continuum. Several technical challenges exist for (at best) inferring bacterial transmission and for (at minimum) understanding the point of flux between vulnerable adults, care and healthcare staff and their immediate environments. First, as transmission of the strain causing infection may have happened long before infection, long-term studies across the care continuum are required. Second, identifying strains associated with clinical infection in gut microbiota requires identifying a single strain in a complex microbial sample and distinguishing between bacteria of the same species contained within a single sample. Third, to infer transmission between two sources, a meaningful distinction must be made between isolates that are derived from the same source and those derived from a different source^12^. Here, we will use the classical microbiological typing terminology to refer to bacteria that are demonstrably different and indistinguishable at the level of genomic resolution we have achieved^13^.

The Tracking Antimicrobial Resistance Across Care Settings (TRACS) Liverpool programme aims to investigate the most important time points in an individual’s care journey from admission to hospital through intermediate care to long-term care facilities for acquisition of ESBL-E/CPE, in particular ESBL- and CP-producing *Escherichia coli* (*E. coli*) and *Klebsiella pneumoniae* (*K. pneumoniae*), and establish the most important transmission routes.

### Primary objective

To detect differences in rates of acquisition of ESBL-E/CPE between patients in hospital and residents in care home settings

### Secondary objectives

1. To identify risk factors for acquisition of ESBL-E/CPE in people in health and social care settings, at the level of the health system, the individual, and the pathogen.
2. To establish a surveillance platform for detection and genotyping of disease-causing and colonising ESBL-E and CPE
3. To describe movement of ESBL-E/CPE bacteria, ESBL and carbapenemase genes and mobile genetic elements within and between hospitals and care facilities
4. To identify putative points for intervention focusing on structural barriers to optimal hygiene practices in long-term residents within the health system and residential care

This process began with a formative study, incorporating Patient and Public Engagement and Involvement, which identified system-level challenges, including data fragmentation, inconsistent infection prevention and control adherence, infrastructure barriers, and communication failures during patient transitions^14^. This information aided in the development of robust, comprehensive protocols from patient^15^ and environment^16^ sampling through to sequence analysis for the targeted genomic surveillance^13^ of ESBL-E and CPE in care settings. Here, we detail the TRACS-Liverpool participant recruitment and microbial surveillance framework with the aim that the protocols used in this programme can be replicated for similar studies.

## Methods

Our protocol is split into six distinct parts: the study design, sample collection, laboratory workflow, sequence data processing and data analysis. Each part will be essential to carry out a thorough transmission study of this scale and requires a range of skills and expertise, including nurses, clinicians, microbiologists and epidemiologists.

### Study design

#### Site Selection

Three different types of healthcare settings will be selected to represent the continuum of care in Merseyside, UK (Figure 1). Selection criteria will include diversity of patient and resident demographics, variation in infrastructure, feasibility of repeated access, and willingness of sites to participate. Settings will include elderly care units within two urban hospitals (one low-acuity care of the elderly ward and one acute frailty unit), one intermediate care facility, and two long-term care homes. These sites will represent differing staff-to-patient or resident ratios, room layouts, and hygiene and sanitation infrastructure. The study will include sites across three NHS trusts and two privately run care homes, reflecting the fragmented nature of elderly care provision in the UK.

**Figure 1.**
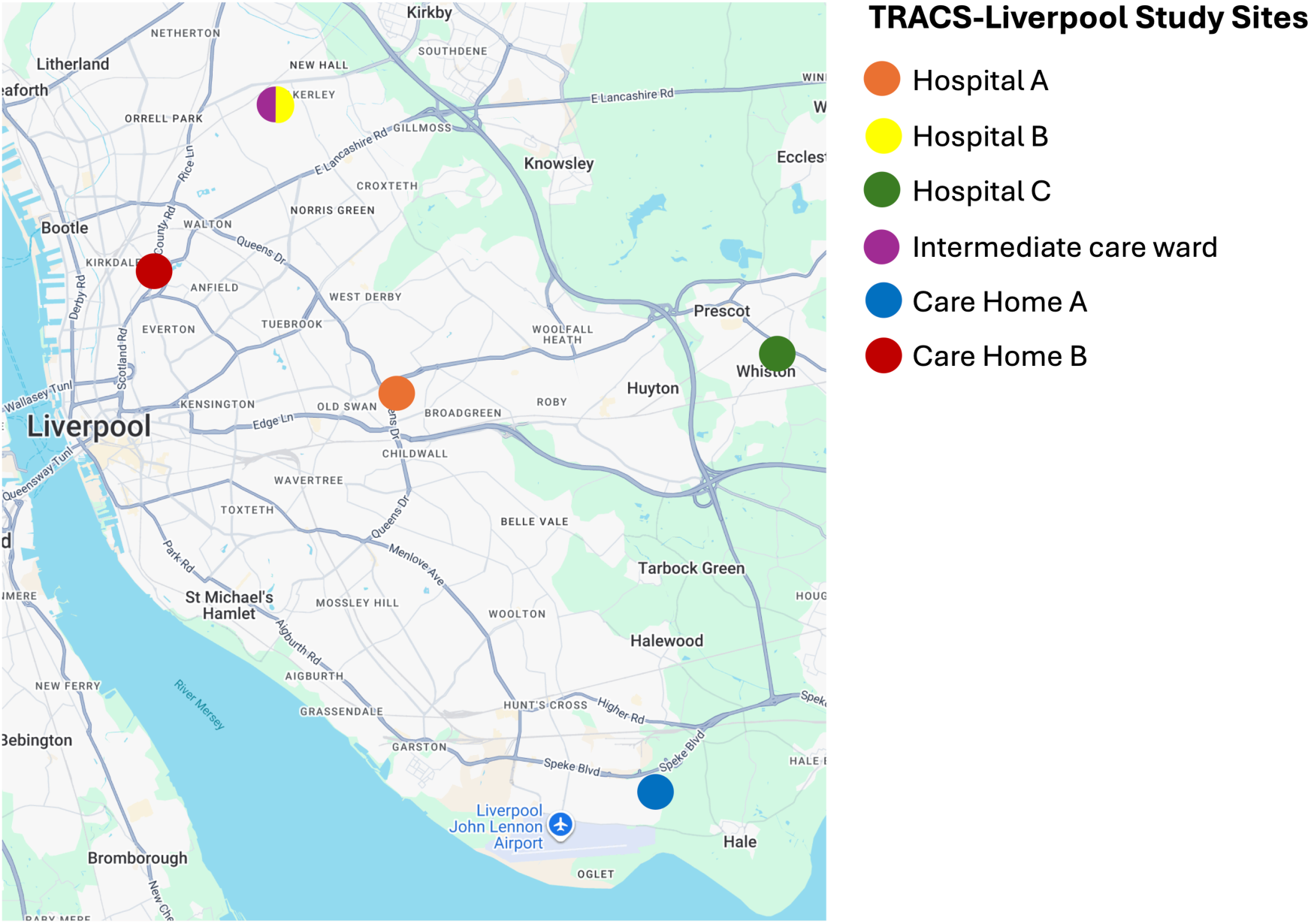
Hospitals and care facilities around Liverpool where environmental and stool/rectal swab samples were collected during the TRACS study. Adapted from Google Maps 2026.

#### Sampling procedure

The study will have a longitudinal design and will span 18 months, incorporating repeated sampling cycles every 12 weeks, rotating across participating sites (Figure 2). Each cycle will consist of a two-week data and sample collection window within the selected study site. During each cycle, stool or rectal swab samples will be collected from patients and residents, and the hands of the staff and the study site environment will be swabbed, according to the sampling schedule outlined in Table 1. This design will allow for the identification of participant demographic and clinical characteristics, description of bacterial colonisation dynamics at individual and facility levels, characterisation of environmental contamination patterns, and assessment of bacterial genomic changes over time.

**Figure 2.**
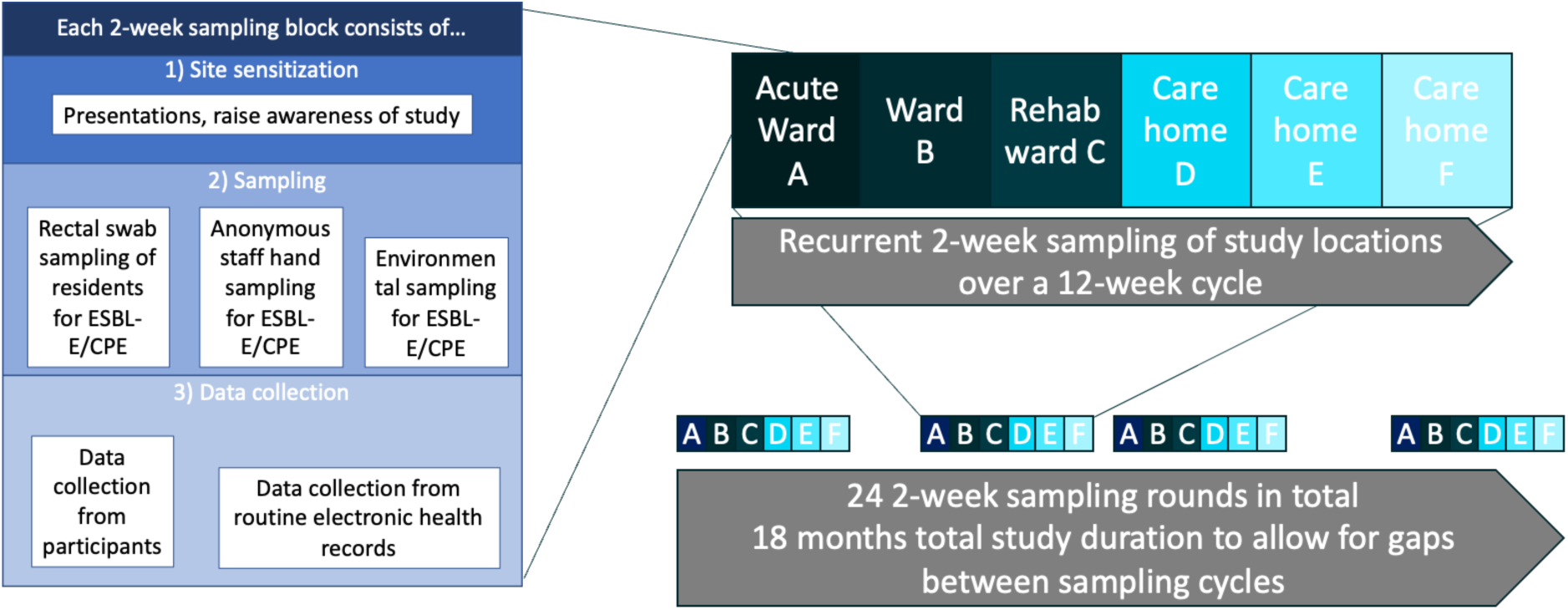
Sampling framework of TRACS-Liverpool study.

**Table 1:**
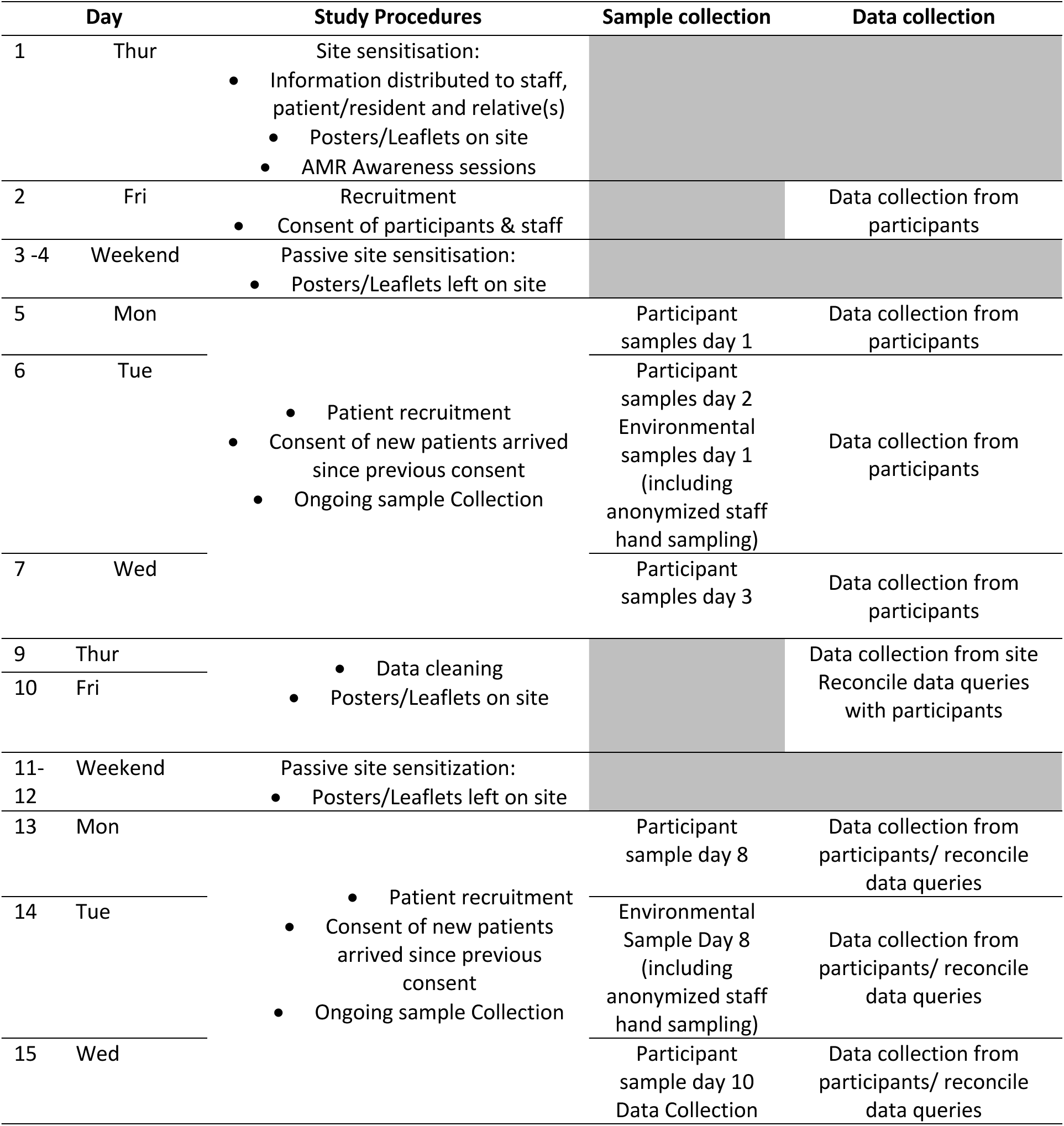
Sampling schedule.

#### Intensive longitudinal sampling

A nested longitudinal sub-study will be conducted, incorporating more frequent longitudinal sampling outside of the two-week sampling windows, to quantify bacterial turnover and flux using high-frequency sampling. Although this approach will not fully resolve all transmission networks, it will allow inference of bacterial turnover over time and will enable improved parameterisation of transmission models.

#### Site set up

Care home managers who participated in the preliminary TRACS qualitative study^14^ will be contacted, alongside additional suitable care homes identified through targeted outreach. Lists of eligible homes will be generated using Care Quality Commission (CQC) and local council data.

It is anticipated that some care homes may continue to face operational pressures related to the COVID-19 pandemic, including staffing shortages and inspection demands, which may affect willingness to participate. NHS hospital and intermediate care sites will be identified through NHS collaborators within the research team and through individual NHS trust research offices.

Sites will be contacted by telephone or email, and each site will be visited at least twice prior to the commencement of sampling. During these visits, site maps for environmental sampling will be developed collaboratively with staff to facilitate engagement and sensitisation. This preparatory work will support successful recruitment and sample collection once the study begins.

### Sample collection

#### Participant Inclusion, Exclusion and Recruitment

Residents and staff will be recruited to the study. Eligible participants in the patient or resident arm will be adults aged 18 years or older residing in, or receiving care from, selected facilities. Exclusion criteria will include patients or residents receiving palliative care where active treatment has been withdrawn.

Potential participants will be screened for eligibility and provided with study information materials, with adequate time to consider participation. Written or online informed consent will be obtained. Where participants lack capacity to consent, procedures will follow the UK Mental Capacity Act (2005), involving consultation with a personal consultee. Participants without an identifiable consultee will not be recruited. It is recognised that those lacking capacity to consent are under-represented in research. Work is being undertaken to improve consultees understanding about health research and their role as consultees. For this study, we will host a nested feasibility trial testing the usefulness of a decision aide for consultees^17^.

Staff members present in the study sampling location during the sampling period will be eligible for recruitment to the study. No personal data will be recorded, and hand sampling will be the only samples collected. It will not be possible to link samples to individuals. For this reason, verbal consent (following the Health Research Authority consent guidance) is considered sufficient.

During site sensitisation activities, staff recruitment and sampling procedures will be explained, and participant information sheets will be distributed. Posters describing the study will be displayed in sampling locations. Staff will have the opportunity to ask questions throughout the study, and those who agree to participate will provide verbal consent prior to hand sampling.

#### Data Collection

Data will be collected using REDCap (Research Electronic Data Capture) tools ^18^ on tablet devices and uploaded to secure servers hosted at the Liverpool School of Tropical Medicine (LSTM). To contextualise microbiological findings, metadata will be collected, including demographic characteristics, healthcare exposures, clinical variables, and movement across care settings. No identifiable data will be collected from staff participants.

#### Sampling Procedures

Stool or rectal swab samples will be collected from patients/residents per Table 1, on designated collection days from all consented participants at each study location. Participants in the main study will not be followed up after discharge or beyond the two-week sampling period. Residents present across multiple sampling periods (e.g. long-term care home residents) will be resampled in subsequent periods like any other consenting resident. Table 1 reflects the maximum sample size assuming recruitment by day 1 and continuous residency.

For the intensive longitudinal sub-study, participants will be additionally sampled on days 14 (±3d), 21 (±3d), 28 (±3d), 56 (±7d), 84 (±7d), 112 (±7d), 140 (±7d), and 168 (±7d). Sampling will occur weekly for the first four weeks, then monthly up to six months. Post-discharge samples can be self-collected and posted to the laboratory.

On resident sampling days, all consenting residents will provide stool (or rectal swab if stool is unavailable). On environmental swabbing days, staff hand swabs and environmental swabs of the following sites in each care setting will be collected:

- Toilet seats and bowls
- Sink taps, outlets, basins, and drains
- Shower rooms, including shower heads and drains
- Door handles, nurses’ worktops, keyboards, telephones
- Medication preparation surfaces

The same sites will be swabbed at each environmental sampling round. These locations were selected based on a pilot exercise and high-risk areas highlighted by previous studies^19–23^ to enrich for likely carriage sites. Sites will be thoroughly swabbed for 20 seconds with a polyester swab (TS/19-G250, Technical Service Consultants swabs) that will be moistened with sterile PBS directly before use, rotating the swab throughout (an approach adapted from Vurayai *et al.* 2022^24^ and Cocker *et al.* 2023^25^).

### Laboratory workflow

#### Processing of samples

The laboratory protocols were optimised to capture ESBL- and CP-producing *E. coli* and *K. pneumoniae* from stool or rectal swabs^15^ and environmental samples^16^ by testing different pre-enrichment media, incubation times and selective plating for the recovery of ESBL-E. Membrane Lactose Glucuronide Agar (MLGA) and Simmons Citrate Agar with Inositol (SCAI), both supplemented with cefotaxime, were selected as the media to select for potential ESBL-producing *E. coli* and *K. pneumoniae* species respectively^6^.

#### Media preparation

Nutrient broths, Buffered Peptone Water (BPW, CM1049B, Thermo Fisher Scientific), Tryptic Soy Broth (TSB, 211 825, BD Biosciences) and solid culture media, MLGA (CM1031B, Thermo Fisher Scientific) and SCAI (CM0155B, Thermo Fisher Scientific, I5125, Merck) will be prepared according to manufacturer’s instructions. Agar plates will be supplemented with 1 µg/mL cefotaxime (sc-202989, Santa Cruz Biotechnology) to select for potential ESBL-producers (EUCAST 2025). For long-term storage of broths post enrichment, a cryopreservation solution will be prepared by dissolving 18.75% w/v maltodextrin (419 699, Merck) and 6.25% w/v trehalose dihydrate (90 210, Merck) into PBS and filter-sterilizing, as developed in Burz et al. 2019 ^26^, with the addition of glycerol to a final concentration of 10% in the cryopreservation solution. This solution increases long-term viability of cells compared to glycerol-only storage solutions.

#### Swab processing and pre-enrichment

The final optimised laboratory workflow is shown in Figure 3. All samples will be transported to the laboratory and processed immediately or stored at 4°C and processed within 24 hours of the sample being received in the laboratory. Rectal swabs (552C, Sterilin), staff hand swabs and environmental swabs will be directly transferred to 5 mL Buffered Peptone Water (BPW). For stool samples, a small amount will be transferred directly to BPW using a 10 µL culture loop. Stool samples and rectal swabs will be incubated for 4 hours, and environmental swabs incubated overnight (16-18 hours) both at 37°C, 220 rpm. Following pre-enrichment, samples will be plated onto both MLGA and SCAI supplemented with 1 µg/mL cefotaxime and incubated overnight at 37°C. 500 µL of the remaining pre-enriched culture will be mixed 1:1 with cryopreservation solution and stored at -80°C.

**Figure 3.**
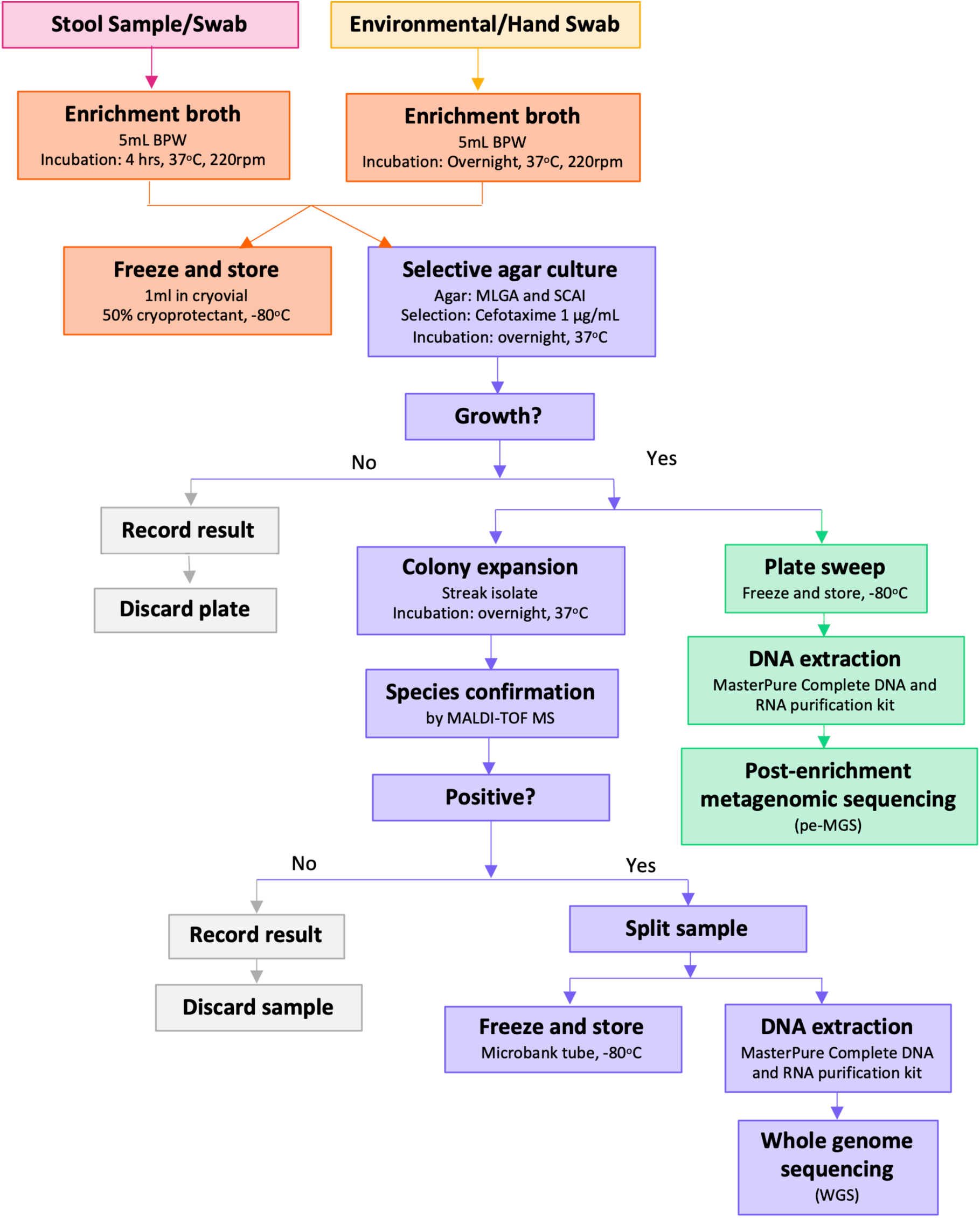
Laboratory workflow for TRACS-Liverpool study from receipt of the sample through to whole genome sequencing. BPW: Buffered Peptone Water

#### Selective agar and interpretation of results

Selective agar resulting in colony growth after plating out pre-enriched samples (termed here “original growth plate”) will be evaluated according to the agar manufacturer’s instructions. Colonies that are either green, blue or yellow on MLGA and yellow on SCAI were selected as potential *E. coli* and *K. pneumoniae*.

#### Species confirmation

A single representative colony for each presumed positive sample will be picked from the original growth plate using a 1 µL culture loop and restreaked onto MLGA (termed here the “restreak plates”) and grown at 37°C overnight. Presumptive positive colonies will undergo species confirmation testing by matrix-assisted laser desorption/ionisation time-of-flight mass spectrometry (MALDI-TOF MS). A single colony will be picked from the restreak plate with a wooden toothpick and applied to the MALDI-TOF target plate. Samples will be prepared by the extended direct transfer method (Bruker) and tested on the MALDI Biotyper Sirius (Bruker). The best match for species identity will be recorded for each colony tested.

For confirmed positive colonies, the remaining growth on the restreak plate will be divided in two, washed in 500 µL sterile PBS (10010023, Thermo Fisher Scientific) and pelleted by centrifugation. One pellet will be used for long-term storage, and the other pellet will be used for DNA extraction prior to sequencing. For long-term storage, the pellet will be resuspended in a Microbank tube (PL.170C, Pro-lab diagnostics) and stored at -80°C. The remaining pellet will be resuspended in 300 µL of tissue and cell lysis solution (MTC096H, Biosearch Technologies) and stored at -80°C prior to DNA extraction.

#### Plate sweeps

All the remaining colonies from the original growth plates will be collected using a 10 µL culture loop, inoculated into a Microbank tube and stored at -80°C prior to DNA extraction. 1 µL of Microbank solution will be diluted in 100µL PBS and plated onto LB agar (L2897, Merck) to provide the input for DNA extraction.

#### Processing of additional samples

To provide additional genomic context, a representative, contemporary collection of *E. coli* and *K. pneumoniae* blood culture isolates from Liverpool Clinical Laboratories and Whiston Pathology Laboratory will be sequenced alongside TRACS samples. We will aim to sequence all available *E. coli* and *K. pneumoniae* isolates from both laboratories, both phenotypically ESBL and non-ESBL producing; for LCL we will aim to sequence all available isolates from 2020-2024; for Whiston (where isolates are stored for a year) we will aim to sample all isolates from 2023, up to a target of 1100 isolates. This number is determined by logistics (primarilily budget available for sequencing). If there are more isolates than 1100, we will aim to sample all ESBL-producing isolates and a random subset of non-ESBL producing isolates up to our target. Isolates will be recovered from frozen stocks either by streaking a bead onto blood agar, or streaking onto nutrient agar slopes, before transport to our laboratory. Single colonies will be inoculated into 1 mL BPW and cultured overnight at 37°C and shaking. These cultures will be extracted using the MagMAX Microbiome Ultra Nucleic Acid Isolation Kit (Thermo Fisher Scientific), following the manufacturer’s guidance for bacterial culture extraction, omitting bead-beating and adding an incubation with RNase A (MRNA092, Biosearch Technologies). Extractions will be automated using the Kingfisher Flex Purification System (Thermo Fisher Scientific) with MagMAX Microbiome Liquid Buccal Flex programme.

#### DNA extraction and sequencing

DNA from confirmed *E. coli* and *K. pneumoniae* isolates and plate sweeps will be extracted using the MasterPure Complete DNA and RNA Purification Kit using the ‘Cell samples’ protocol provided by the manufacturer (MC85200, Biosearch Technologies). Libraries for genomic DNA of all isolates will be constructed using NEB Ultra II custom kit on Agilent Bravo WS automation system and sequenced on the Illumina HiSeq X10 platform (Illumina, San Diego, CA, USA) at the Wellcome Sanger Institute, UK.

### Sequence data processing

#### Bioinformatic analysis of isolates

The short read sequence data will be assembled *de novo* using the nf-co.re/bacass pipline^27^, which uses Unicycler (version 0.5.1). The quality of the sequence data will be assessed using CheckM (version 1.2.4) and QUAST (version 5.3.0), all sequences that meet the following criteria will be included in subsequent analysis:

- Completeness > 90%
- Contamination < 5 %
- Total assembled length > 4Mb and < 6.5Mb
- N50 > 20kb

Bacterial species will be assigned using the Kraken2/ Braken pipeline (version 1.3.0). Multi-locus sequence typing (MLST) will be performed *in silico* with the mlst tool (version 2.23.0) to determine sequence types^14^. We will screen for AMR genes using AMRFinderPlus (version 4.0.23) ^15^. We will then perform pangenome analysis and generate a core gene alignment using panaroo (version 1.3.4)^16^. A core gene maximum likelihood phylogenetic tree will be constructed using IQ-TREE (version 2.3.6) with the GTR+F model^17^. To analyse transmission networks and clustering of isolates, a pairwise SNV distance comparison between each isolate will be created using snp-dists version 0.8.2 (https://github.com/tseemann/snp-dists).

#### Bioinformatic analysis of plate sweeps

Species assignment for the plate sweep sequence data was done using the mSWEEP (v 2.2.0) and mGEMS (v 1.3.3) algorithm^28,29^, using a curated reference database consisting of *E. coli* and *Klebsiella species complex* assemblies. Samples will then be extracted from the relevant read bins using mGEMS. Multi-locus sequence typing will be done using ARIBA (v2.11.1)^30^ with the seven-gene Achtman scheme and a pairwise SNV distance comparison between each sample will be created using our SNV calling workflow^13^.

### Data analysis plan

#### Primary analysis

The analysis will use Markov hidden state transmission models ^31,32^ where recruited participants can, at any time, be in an ESBL-E/CPE colonised or uncolonised state. Starting with a simple model, this approach will allow an estimation of differences in ESBL-E/CPE acquisition and loss between acute care (hospital) and social care settings (i.e. address the primary outcome). It will then be expanded to include covariates that can influence rates of gain/loss and, ultimately, be expanded to a full agent-based model, accounting for within-facility person-to-person transmission, leveraging previous work by our research group and collaborators^25^, and addressing secondary outcomes.

We used a simulation approach to sample size calculation, using R v4.1.1 (R foundation for statistical computing, Vienna, Austria) and the msm v1.6.9 package^32^. Our primary outcome is the difference between ESBL-E/CPE acquisition in hospitals and care homes; we assume that we wish to detect a hazard ratio of at least 2 (i.e. a twofold difference in the instantaneous rate of ESBL acquisition) between these settings. We used routinely collected ESBL/CPE screening swab data from patients admitted to LUHFT critical care areas (where samples are taken on admission then weekly) to estimate baseline ESBL-E/CPE acquisition and loss rates using msm. We then used these rates to generate 1000 simulated studies with a given number of participants, n and simulated sampling of participants as per the sampling schedule above. We accounted for uncertainty in gain and loss parameter values by sampling from the multivariate normal distribution of these parameters for each simulation, with covariance matrices determined by the msm fit. In each study we included one covariate, indicating hospital versus nursing home location, assigning it a hazard ratio of 3.

We then fit a msm multistate model to each simulation. A given simulation was said to have correctly identified an effect of hospitalisation at the 5% level if the 95% confidence interval of the estimated hazard ratio of hospitalisation did not cross 1 (Figure 4). The power to correctly identify this effect for a given n was defined as the proportion of the 1000 simulations which correctly identified an effect of hospitalisation at the 5% level. 155 Participants, fully sampled, will allow detection of a HR of 3 at 80% power (Figure 4).

**Figure 4:**
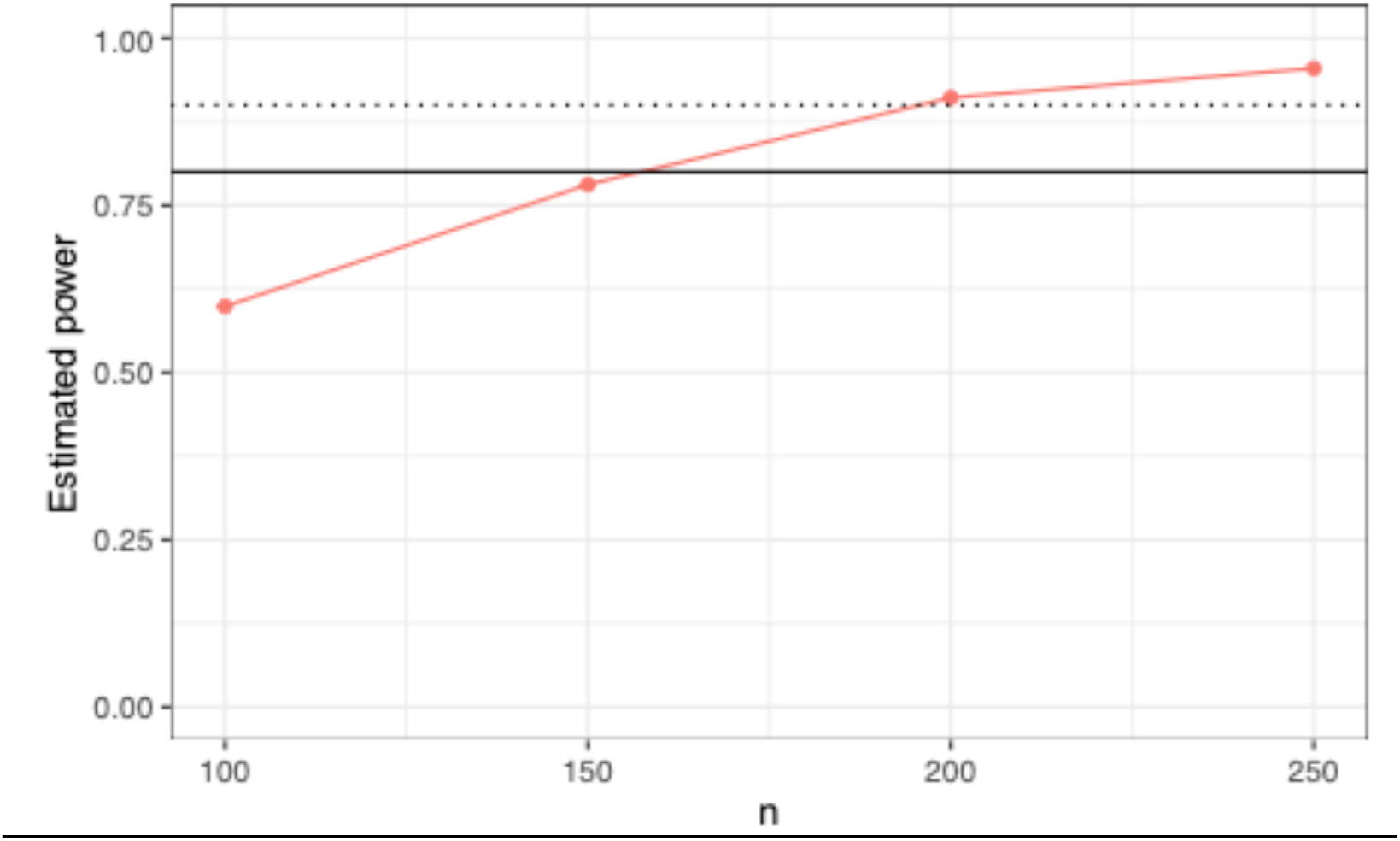
Estimated power for given sample size. The solid line indicates the sample size that gives 80% power, and the dotted line indicates the sample size that gives 90% power.

#### Secondary analyses

Secondary objective 1: Additional covariates will be added into the multistate models to investigate their effect on ESBL-E/CPE acquisition and loss: antimicrobial exposure as a time-varying covariate, spatial proximity to environmental contamination, and force of infection from other colonised individuals on the ward. This latter approach will require generalisation of the multistate modelling framework using the SMC2 algorithm as previously developed by our research group and collaborators for the DRUM (Drivers of resistance in Uganda and Malawi) project^25^.

Secondary objective 2: This analysis will take the form of whole-genome sequencing of cultured isolates followed by assembly (by mapping to a reference) and calculation of single nucleotide variants (SNV) distance between isolates. An empiric SNV distance cut-off will be derived by comparing within- to between- participant isolates and identifying a cut-off that can distinguish the two. This cut-off will then be used to identify putative transmission events. A descriptive analysis of the frequency of transmission between participants and the environment will be presented.

Secondary objective 3: This analysis will take the form of shotgun metagenomic sequencing of bacteria from plate-sweeps, which we will refer to as “post-enrichment metagenomic sequencing” to describe ESBL-E/CPE diversity within participant as they pass along their care journey and/or are exposed to antibiotics. A descriptive analysis of within-participant diversity will be presented.

#### Data management

Data will be entered directly onto tablet devices and will be considered source data. Other source data will be data that are extracted from routine electronic health records. Source data will be stored securely on LSTM servers, with regular backup, managed by the experienced Global Health Trials Unit (GHTU) at LSTM, with SOPs for the use of the system, and audit trial for data changes, using the REDcap platform. Access to the data will be restricted to the data and study team members.

REDcap is a secure, web-based software platform designed to support data capture for research studies, providing 1) an intuitive interface for validated data capture; 2) audit trails for tracking data manipulation and export procedures; 3) automated export procedures for seamless data downloads to common statistical packages; and 4) procedures for data integration and interoperability with external sources.

Participants will be identified by an unambiguous TRACS study code, which will allow identification of all the data for each participant. This pseudonymised data will be linked back to identifiable participant information, with secure maintenance of personal data and the linking study identifier in separate locations using encrypted, password protected storage. Access to personally identifiable data will be limited to the minimum number of individuals necessary for quality control, audit, and analysis. Archiving will be authorised by the Sponsor following submission of the end of study report. All archived documents (source data, CRFs, study databases) will be stored on secure LSTM servers for a minimum of five years following completion of the study.

## Ethics

The collection of clinical samples for the TRACS-Liverpool study was approved by the National Research Ethics Service Greater Manchester South ethics committee (ref: 22/NW/0343). All procedures were conducted in accordance with the UK Policy Framework for Health and Social Care Research. Written informed consent was obtained from participants or consultees, as appropriate. Clinical trial number: not applicable.

## Discussion

Improved epidemiological and laboratory approaches are required to study asymptomatic bacterial transmission across healthcare settings. Our protocol, tailored to this challenge, has several strengths. First, the inclusion of staff, patient, and environmental sampling will provide a holistic understanding of AMR ecology. Second, longitudinal repeated sampling over 18 months will enhance our ability to detect transient colonisation, intermittent shedding, and environmental persistence patterns, perhaps before cases of disease. Third, integration with genomic sequencing will allow for the high-resolution mapping of transmission clusters that would otherwise be missed by conventional phenotyping. Lastly, the rigorous statistical modelling will allow for the detection of differences in acquisition rates between acute care and social care settings and identify risk factors at individual, facility, and pathogen levels.

Several challenges are anticipated during the conduct of this study. Setting up could take longer than expected due to difficulties engaging with NHS trusts and independent care sites, with added complexity arising from recruiting participants lacking capacity and involving consultees. Staffing issues may arise, and many patients, particularly in hospital settings, may be unwilling to participate, since they may already be fatigued by their hospital experience. Logistically, transporting samples to the lab within required timeframes across a busy urban area (with extended journeys), which will limit the amount of dedicated recruitment time, a problem likely to be worsened by limited parking at all hospital sites. Increased transport times are likely to negatively impact the recovery of ESBL/CP-E in the laboratory. This is due to several factors occurring during transit, including desiccation, temperature fluctuations, and the extended period between sample collection and processing, during which the ESBL/CP-E may become unrecoverable.

Despite these limitations, the integrated approach will significantly advance the field by providing a protocol capable of supporting multilevel analysis - clinical, ecological, infrastructural, and genomic. The design is scalable and adaptable, offering a blueprint for regional or national AMR surveillance systems.

## Conclusion

We present a comprehensive and rigorous protocol for surveilling transmission for ESBL/CPE-E across the continuum of care. By incorporating study participant, staff, and environmental sampling with an enriched contextual dataset and genomic analysis pipeline, this protocol provides a holistic framework for studying AMR transmission. This approach enhances the ability to understand acquisition pathways, identify modifiable environmental reservoirs, and to evaluate and guide IPC interventions in both hospitals and long-term care facilities. The protocol described here is readily adaptable to other regions and supports national surveillance efforts that integrate environmental microbiology, infection prevention, and genomic epidemiology.

## Data availability

All the standard operating procedures that will be used in the TRACS-Liverpool study are publicly available on Zenodo: Standard Operating Procedures (SOPs) used for publication: A protocol for the TRACS-Liverpool study, tracking transmission of extended-spectrum beta-lactamase producing Enterobacterales across health and social care settings in the United Kingdom, 10.5281/zenodo.19571592.

This project contains the following data:

- TRACS_SOP_001_ Environmental_swab_sampling_V4.pdf
- TRACS_SOP_002_ Staff_hand_sampling_V1.pdf
- TRACS_SOP_003_ Stool_Rectal_swab_sampling_V1.pdf
- TRACS_SOP_004_Environmental_sample_processing_V3.pdf
- TRACS_SOP_005_Stool_sample_processing_V3.pdf
- TRACS_SOP_006_MasterPure_DNA_extraction_V2.pdf

All the case report forms that will be used in the TRACS-Liverpool study on the REDCap digital platform are publicly available on Zenodo: Case Report Forms (CRFs) used for publication: A protocol for the TRACS-Liverpool study, tracking transmission of extended-spectrum beta-lactamase producing Enterobacterales across health and social care settings in the United Kingdom, 10.5281/zenodo.19605278.

This project contains the following data:

- TRACS_REDCap_CRFs.pdf

## References

1. Naghavi, M. et al. Global burden of bacterial antimicrobial resistance 1990–2021: a systematic analysis with forecasts to 2050. The Lancet 404, 1199–1226 (2024).

2. Javadi, A. et al. Qualification Study of Two Genomic DNA Extraction Methods in Different Clinical Samples. Tanaffos 13, 41 (2014).

3. Cocker, D. et al. Healthcare as a driver, reservoir and amplifier of antimicrobial resistance: opportunities for interventions. Nature Reviews Microbiology 2024 22:10 22, 636–649 (2024).

4. Sati, H. et al. The WHO Bacterial Priority Pathogens List 2024: a prioritisation study to guide research, development, and public health strategies against antimicrobial resistance. Lancet Infect. Dis. 25, 1033–1043 (2025).

5. Karthika, M., Abraham, J., Kodali, P. B. & Mathews, E. Emerging Trends of Chronic Diseases and Their Care Among Older Persons Globally. Handbook of Aging, Health and Public Policy 1–24 (2023) doi:10.1007/978-981-16-1914-4_198-1.

6. Older Person Care Homes Liverpool. https://www.carehome.co.uk/care_search_results.cfm/searchtown/Liverpool/searchchtype/old-age.

7. Lowe, C. et al. Outbreak of Extended-Spectrum β-Lactamase–producing Klebsiella oxytoca Infections Associated with Contaminated Handwashing Sinks. Emerg. Infect. Dis. 18, 1242 (2012).

8. Mitchell, B. G., Dancer, S. J., Anderson, M. & Dehn, E. Risk of organism acquisition from prior room occupants: a systematic review and meta-analysis. Journal of Hospital Infection 91, 211–217 (2015).

9. Decraene, V. et al. A large, refractory nosocomial outbreak of klebsiella pneumoniae carbapenemase-producing Escherichia coli demonstrates carbapenemase gene outbreaks involving sink sites require novel approaches to infection control. Antimicrob. Agents Chemother. 62, (2018).

10. Karanika, S., Karantanos, T., Arvanitis, M., Grigoras, C. & Mylonakis, E. Fecal Colonization With Extended-spectrum Beta-lactamase–Producing Enterobacteriaceae and Risk Factors Among Healthy Individuals: A Systematic Review and Metaanalysis. Clinical Infectious Diseases 63, 310–318 (2016).

11. Thompson, D. C. et al. The Impact of COVID-19 Pandemic on Long-Term Care Facilities Worldwide: An Overview on International Issues. Biomed Res. Int. 2020, 8870249 (2020).

12. Balloux, F. et al. From Theory to Practice: Translating Whole-Genome Sequencing (WGS) into the Clinic. Trends Microbiol. 26, 1035–1048 (2018).

13. Gallichan, S. et al. A more complete picture: Capturing single nucleotide variant diversity in extended-spectrum beta-lactamase producing Escherichia coli using post-enrichment metagenomics. medRxiv 2025.11.12.25340063 (2025) doi:10.1101/2025.11.12.25340063.

14. Alhassan, Y. et al. Health system drivers of antimicrobial resistance: a qualitative exploration of implications for infection prevention and control in hospitals and long-term care facilities in Merseyside. Journal of Hospital Infection 166, 12–20 (2025).

15. Gallichan, S. et al. Optimized methods for the targeted surveillance of extended-spectrum beta-lactamase-producing Escherichia coli in human stool. Microbiol. Spectr. 13, (2025).

16. Picton-Barlow, E. et al. An evaluation of screening methods for the detection of extended-spectrum beta-lactamase-producing Escherichia coli and Klebsiella pneumoniae in environmental samples from healthcare settings. J. Appl. Microbiol. 136, 295 (2025).

17. Shepherd, V. et al. Feasibility of a Study Within a Trial to evaluate a decision support intervention for families deciding about research on behalf of adults lacking capacity to consent (CONSULT SWAT). Trials 2025 26:1 26, 313- (2025).

18. Harris, P. A. et al. Research electronic data capture (REDCap)—A metadata-driven methodology and workflow process for providing translational research informatics support. J. Biomed. Inform. 42, 377–381 (2009).

19. Decraene, V. et al. A large, refractory nosocomial outbreak of klebsiella pneumoniae carbapenemase-producing Escherichia coli demonstrates carbapenemase gene outbreaks involving sink sites require novel approaches to infection control. Antimicrob. Agents Chemother. 62, (2018).

20. Christoff, A. P. et al. One year cross-sectional study in adult and neonatal intensive care units reveals the bacterial and antimicrobial resistance genes profiles in patients and hospital surfaces. PLoS One 15, e0234127 (2020).

21. Jamal, A. J. et al. Carbapenemase-producing Enterobacterales in hospital drains in Southern Ontario, Canada. Journal of Hospital Infection 106, 820–827 (2020).

22. Glowicz, J. B. et al. SHEA/IDSA/APIC Practice Recommendation: Strategies to prevent healthcare-associated infections through hand hygiene: 2022 Update. Infect. Control Hosp. Epidemiol. 44, 355–376 (2023).

23. Kuczewski, E. et al. Bacterial Cross-Transmission between Inanimate Surfaces and Patients in Intensive Care Units under Real-World Conditions: A Repeated Cross-Sectional Study. International Journal of Environmental Research and Public Health 2022, Vol. 19, Page 9401 19, 9401 (2022).

24. Vurayai, M. et al. Characterizing the bioburden of ESBL-producing organisms in a neonatal unit using chromogenic culture media: a feasible and efficient environmental sampling method. Antimicrob. Resist. Infect. Control 11, 14 (2022).

25. Cocker, D. et al. Drivers of Resistance in Uganda and Malawi (DRUM): a protocol for the evaluation of One-Health drivers of Extended Spectrum Beta Lactamase (ESBL) resistance in Low-Middle Income Countries (LMICs). Wellcome Open Res. 7, 55 (2023).

26. Burz, S. D. et al. A Guide for Ex Vivo Handling and Storage of Stool Samples Intended for Fecal Microbiota Transplantation. Sci. Rep. 9, (2019).

27. Ewels, P. A. et al. The nf-core framework for community-curated bioinformatics pipelines. Nature Biotechnology 2020 38:3 38, 276–278 (2020).

28. Mäklin, T. et al. Bacterial genomic epidemiology with mixed samples. Microb. Genom. 7, 000691 (2021).

29. Maklin, T. et al. High-resolution sweep metagenomics using fast probabilistic inference [version 1; peer review: 1 approved, 1 approved with reservations]. Wellcome Open Res. 5, 1–20 (2020).

30. Hunt, M. et al. ARIBA: rapid antimicrobial resistance genotyping directly from sequencing reads. Microb. Genom. 3, (2017).

31. Lewis, J. M. et al. Colonization dynamics of extended-spectrum beta-lactamase-producing Enterobacterales in the gut of Malawian adults. Nature Microbiology 2022 7:10 7, 1593–1604 (2022).

32. Jackson, C. H. Multi-State Models for Panel Data: The msm Package for R. J. Stat. Softw. 38, 1–28 (2011).

